# TESTING INFORMED SIR BASED EPIDEMIOLOGICAL MODEL FOR COVID-19 IN LUXEMBOURG

**DOI:** 10.1101/2020.07.21.20159046

**Authors:** Thomas Sauter, Maria Pires Pacheco

## Abstract

The interpretation of the number of COVID-19 cases and deaths in a country or region is strongly dependent on the number of performed tests. We developed a novel SIR based epidemiological model (*SIVRT*) which allows the country-specific integration of testing information and other available data. The model thereby enables a dynamic inspection of the pandemic and allows estimating key figures, like the number of overall detected and undetected COVID-19 cases and the infection fatality rate. As proof of concept, the novel *SIVRT* model was used to simulate the first phase of the pandemic in Luxembourg. An overall number of infections of 13.000 and an infection fatality rate of 1,3% was estimated, which is in concordance with data from population-wide testing. Furthermore based on the data as of end of May 2020 and assuming a partial deconfinement, an increase of cases is predicted from mid of July 2020 on. This is consistent with the current observed rise and shows the predictive potential of the novel *SIVRT* model.

## INTRODUCTION

The pandemic disease COVID-19, caused by the coronavirus SARS-CoV-2, became in a few months one of the leading causes of death worldwide with now over 580.000 fatalities and 13 million reported cases (Dong et al., 2020; *John Hopkins Dashboard July 16, 2020*, n.d.). The total number of cases and recovered patients is unknown as a fraction of the virus carriers do only show mild or no symptoms and hence, escape any diagnostics or could not get tested especially at the onset of the crisis due to a lack of infrastructure and test material. To a lesser extent, the number of cases is likely to be underestimated in countries that did not count deaths outside care facilities. Whereas other countries like Belgium included every fatality that has been tested positive for the virus regardless of the cause of death (https://www.politico.eu/article/why-is-belgiums-death-toll-so-high/). The lack of consistency among testing strategies and case counts prevents the reliable and comparable calculation of simple measures, such as the infection fatality rate (IFR) or the effective reproduction number *Rt_eff*, which is required to better assess the virulence and spread of the disease. A unified large-scale testing strategy and a more rigorous integration of the testing information would enable more precise political decisions on measures beyond following the all-or-nothing example of China, which imposed a lockdown on their population to avoid a breakdown of the healthcare system due to a saturation of ICU beds by COVID-19 patients. Among the first affected countries by the virus, only the ones that had experienced the previous MERS-CoV outbreak such as South Korea and Singapore and hence had mitigation strategies in place or had established large test infrastructures like Iceland could avoid strict containment strategies. Other countries like Sweden and England attempted to find a balance between lockdown and uncontrolled spread, by slowing the contagion to protect the healthcare system and the elderly people without facing the economic harm caused by a full lockdown.

In Luxembourg, the first case and death were reported on the 29^th^ of February and the 13^th^ of March 2020, respectively. On the 16^th^ of March, schools were closed and all non-crucial workers shifted to remote work or were furloughed. Two days later, the state of emergency was declared, in-person gatherings were prohibited, and restaurants and bars were closed. Over 70.000 workers were furloughed and 30.000 more took a leave for family reasons to homeschool their children. The costs of these measures in the early phase of the pandemic (until April 6th, 2020) were already estimated to be over 150 million euros and 226 million, respectively. To date, 111 people died of COVID-19 with around 4000 cases being diagnosed. A third of these cases required medical attention. Initially, Luxembourg performed diagnostic tests based on swabs in the nose or mouth to detect the presence of the virus by PCRs, thereby prioritizing travelers, and their close family circle. Later, Luxembourg could ramp up their testing facility and could test citizen and cross-border workers having a medical recipe. From the 27^th^ of May to the 28^th^ of July 2020, 17 drive-in stations were put in place that allowed 20.000 tests per day. The aim was to test the whole population in function of the degree of exposure to the virus.In parallel, within the CON-VINCE study serologic tests were performed to assess the presence of IgG and IgA in the plasma, as well as nose and mouth swabs on a random set of 1.800 habitants to assess the spread of the disease in the Luxembourgish population. Around 1,9% of the samples had antibodies and 5 people tested positive, indicating that Luxembourg was far away from herd immunity (Snoeck et al., 2020).

Epidemic models such as compartment models have proven to be a useful tool in other outbreaks to access the efficiency of mitigation strategies and to plan the timing and strength of interventions. More specifically, *SIR* (Susceptible, Infected and Removed) and *SEIR* (Susceptible, Exposed, Infected, and Removed) models, formulated as Ordinary Differential Equations (ODE), allow determining when social distancing, hand washing, testing, and voluntary remote working should be sufficient to prevent an exponential growth of the cases and when a significant portion of the population has to return into lockdown (Song et al., 2020; Tang et al., 2020; Wangping et al., 2020; Yang et al., 2020). Adaptations and extensions of *SIR* and *SEIR* models were published for COVID-19 already (Giordano et al., 2020; Siwiak et al., 2020; Tang et al., 2020). Such models allow describing the dynamics of mutually exclusive states such as Susceptible (S) which for COVID-19 is assumed to be the entire population of a country, a region or city, the number of Infected (*I*) and Removed (*R*) that often combines (deaths and recovered), as well as the number of Exposed (*E*) for *SEIR* models. The variables *I* and *R*, are often unknown as the number of cases and announced recovered patients only accounts for a fraction of the real values that is dependent on the testing performed within a country. Therefore the numbers of Susceptible and Exposed that equals the total population minus Infected and Removed in *SEIR* models are also undetermined. Several studies extended the number of considered states in such models to further differentiate between detected and undetected cases (Susceptible (*S*), Infected (*I*), Diagnosed (*D*), Ailing (*A*), Recognized (*R*), threatened (*T*), Healed (*H*) and Extinct (*E*)), (Gaeta, 2020; Giordano et al., 2020) or took the severity of the disease into account in relation to the age of the infected person (Balabdaoui & Mohr, 2020; Wu et al., 2020). However, with an increase in the number of states and parameters describing the transitions between these states, more data is required to calibrate the model, i.e. to estimate the model parameters. Roda et al (Roda et al., 2020) showed that an *SIR* model seems to better represent data obtained from case reports than *SEIR* models. Notably, *SIR* models captured a link between the transmission rate β and the case-infection ratio that was missed by *SEIR* models. The underestimation of the Infected, Deaths, and Removed, due to not considering country specific testing information, causes *SIR* models to predict IFR and effective reproduction number (*Rt_eff*) values that vary drastically across countries with different testing and might often be overestimated. To overcome this issue, we propose an extended SIR model (*SIVRT*) which is informed by the number of performed tests and also takes the number of hospitalizations into account to parametrize the model. This allows for a better prediction of the evolution of the disease and the estimation of key pandemic parameters, as well as the analysis of different deconfinement and testing strategies.

## METHODS

### Model

The novel *SIVRT* model (Figure 1 & Appendix 1) is formulated with ordinary differential equations and consists of 9 states: Susceptible (*S*), Infected (*I*), Recovered (R), seVere cases (*S*), Death cases (*D*), Detected Infected (*I*_*D*_), Detected seVere cases (*V*_*D*_), Detected Recovered (*R*_*D*_) and Detected Death cases (*D*_*D*_). Note that the model distinguishes between non-detected and detected values. Relevant summary outputs are reported in terms of overall *Cases* (CasesAct = I + ID + V + VD), *Recovered* (RCum = R + RD) and *Dead* (DeadCum = DeadD + Dead). All states are given per day, starting on Feb 28, 2020, resembling Day 0 of the simulations.

**Figure 1:**
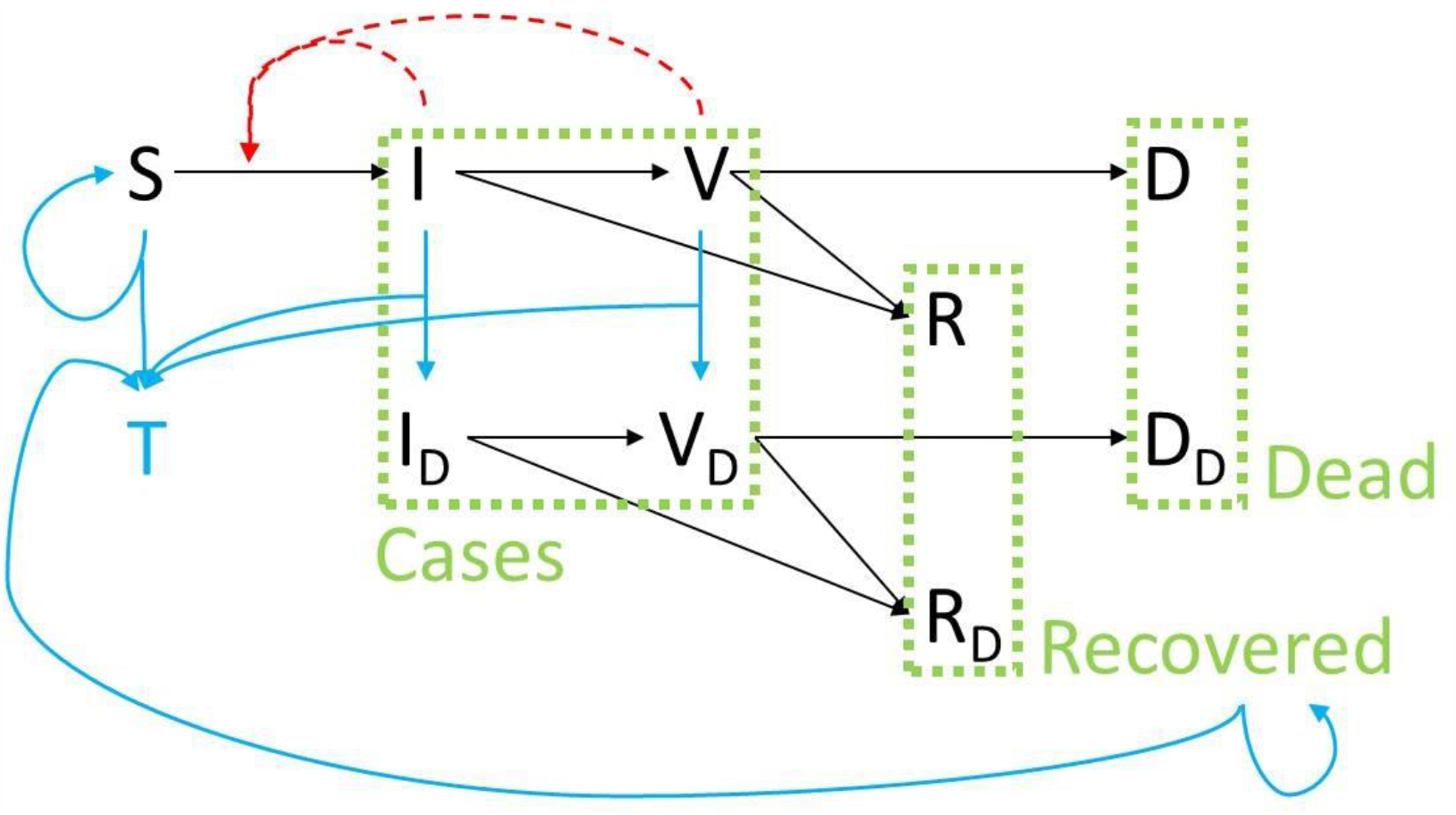
*SIVRT* model structure: Starting from the susceptible population (*S*), individuals might become infected (*I*) and subsequently either recover (*R*) or develop a severe form of the disease (*S*). Severe cases either recover (*R*) or die (*D*). A second parallel layer describes the same path for the detected number of infected (*I*_*D*_), severe cases (*V*_*D*_), recovered (*R*_*D*_), and death cases (*D*_*D*_). This second layer is reached by testing (blue arrows) from the first undetected layer. The overall number of tests is denoted by *T*. Infection is assumed to be only transmitted by the undetected infected and severe cases (red arrow). Relevant summary outputs are reported in terms of overall *Cases, Recovered* and *Dead* (dashed green boxes).

The number of daily tests (*T*) is also modelled as a state, but its dynamics are fitted by a polynomial of 4^^th^^ order using the *polyfit* function in Matlab prior to the model simulation. The derivative of the fitted polynomial is then included as balance equation for *T*.The model contains 8 parameters (listed in Appendix 1, with a representative set of parameter values) describing the transitions between the states, as well as 1 additional parameter per event. An event thereby describes the change of the infection rate constant (kSI) when changing the lockdown strategy. For Luxembourg, the lockdown began on Day 17 of the simulations (resembling March 16, 2020).

Concerning the model structure and parameter values, the following rationales and assumptions have been applied: The Susceptible *S* are infected by non-detected cases with a rate constant kSI and under the consideration of a Monod type saturation for *S*. It is supposed that detected cases are not contributing to the spreading of disease because of quarantine measures. This could be adjusted in other model versions if appropriate. After a lockdown event (one for Luxembourg on March 16 = Day 17) the value of rate constant kSI can be decreased during the fitting by a factor ranging from 0% to 100% of the starting value. Infected (*I)* and DetectedInfected (*I*_*D*_) can further transition to Recovered (*R)* or severe cases (*V)* and Detected Recovered (*R*_*D*_) or Detected seVere cases (*V*_*D*_), respectively. Severe cases (*V)* can evolve to Recovered (*R)* or Death cases (*D)* and Detected Severe cases (*V*_*D*_) to Detected Recovered (*R*_*D*_) and Detected Death cases (*D*_*D*_). All these transitions are modeled with first-order laws with rate constants kIR, kIV, kVR, kVD, kIDR, kIDVD, kVDR, and kVDD, respectively. The rate constants for detected and non-detected states are assumed to be equal. Regarding the testing, it is presumed that (i) Severe cases S are tested with high probability compared to asymptomatic cases, as it is more likely that the severe cases will be spotted within the population; (ii) Susceptible S and Recovered people *RCum* (=R+RD) are tested with the same probability. Testing of Severe cases S is modelled as a first-order term as well (kVVD), and the remaining performed tests are distributed among Infected (*I)* and the sum of Susceptible and Recovered (*S+RCum*). The ratio between these two groups is adjusted with parameter kTIvsS which is also subjected to optimization.

### Data

For Luxembourg data on people tested positive, death cases, hospitalisation and performed tests were obtained from the website of the Luxembourgish government https://coronavirus.gouvernement.lu/en.html and are summarized in Appendix 2.

### Model Fitting

The model was implemented in the IQM toolbox (Sunnåker & Schmidt, 2016) within the Matlab framework and is given in Appendix 1. The data integration as well as the estimation of all 9 model parameters was performed within IQM by applying a combined global (*pswamIQM*) and local (*simplexIQM*) optimization. Lower and upper bounds were set as follows: kSI: 1e-06, 2; kmSI: 62611, 187830; kIR: 0, 1; kIV: 0.02, 1; kVR: 0, 1; kVD: 0, 1; kVVD: 0.1, 1; kTIvsS: 2, 200; kSIld: 0.1, 0.8. The optimization was repeated 100 times and successful fits with an *optimalcost* smaller or equal 0.01 were retained for simulation and prediction. Simulation and prediction results are shown as the median of all successful model fits (dashed line), with red solid lines indicating confidence intervals containing 68% of the successful fits.

### Predictions

For the predicted no lockdown at all scenarios (Figure 3), the lockdown event on Day 17 was removed. For the predicted light lockdown scenario (Figure 4), the lockdown event on day 17 was kept, but the infection rate parameter (kSI) was increased by 10% of the difference of its value during the full lockdown compared to its value before the lockdown. For the predicted partial lifting of the lockdown scenario as of end of May (Figure 5), the infection rate parameter (kSI) was increased on Day 85 of the simulation by 20% of the difference of its value during the full lockdown compared to its value before the lockdown. The testing rate was kept constant. For the predicted lifting of the lockdown scenario as of end of May with increased testing approximately matching the Luxembourgish strategy of testing (Figure 6), on Day 85 of the simulation the infection rate parameter (kSI) was set to its value before the lockdown and the testing rate was increased to 5.000 tests per day.

**Figure 2:**
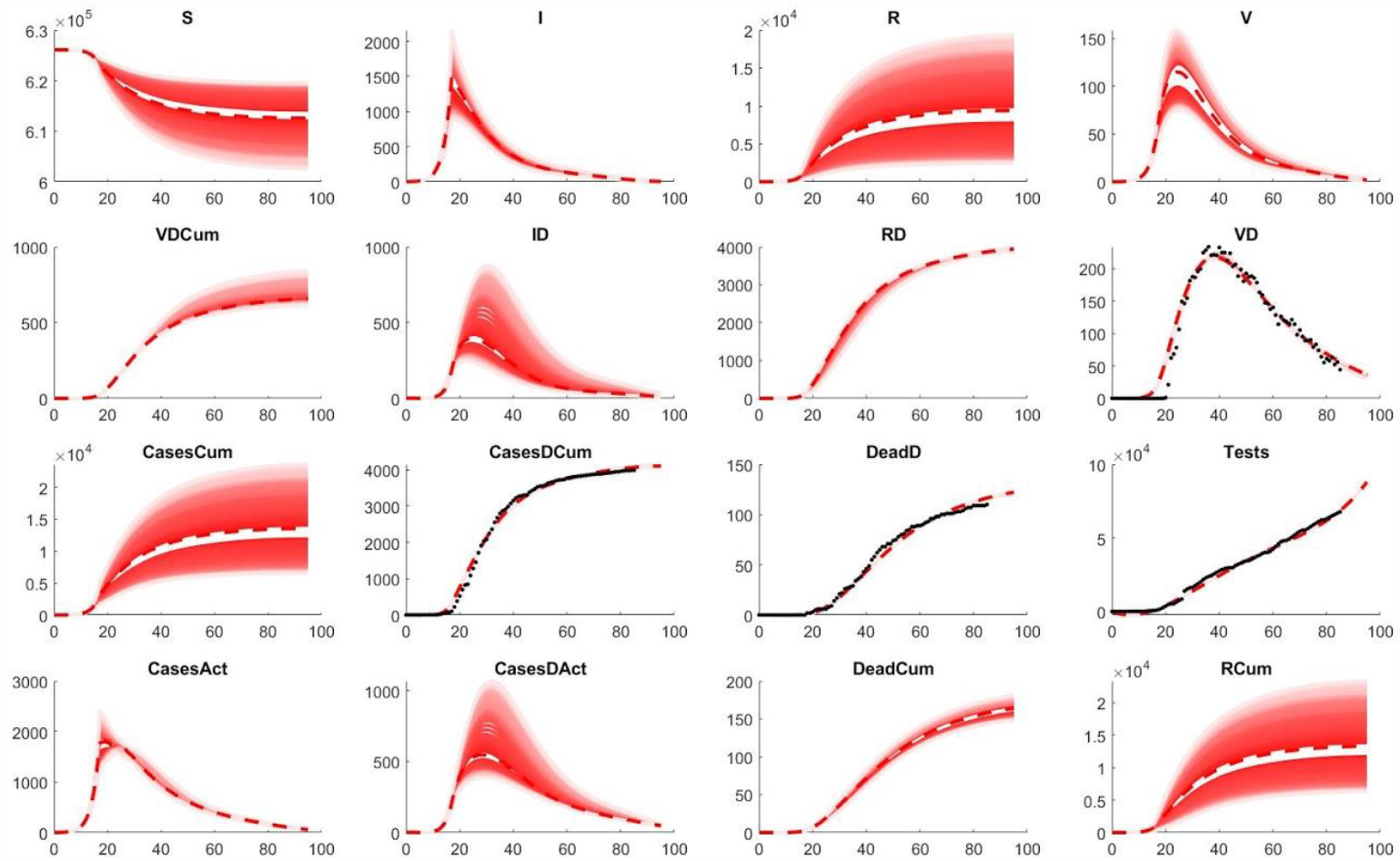
COVID pandemic in Luxembourg. Comparison of observed data and model fit, given in days (x-axis). Black dots represent observed data and red dashed lines represent the median of all successful model fits, with red solid lines fading up to 68% confidence. Subscript *D* indicates detected cases and the ending *Cum* indicates cumulative numbers of the respective variables, e.g. *CasesCum* shows the estimated overall cases in Luxembourg (detected & non-detected).

**Figure 3:**
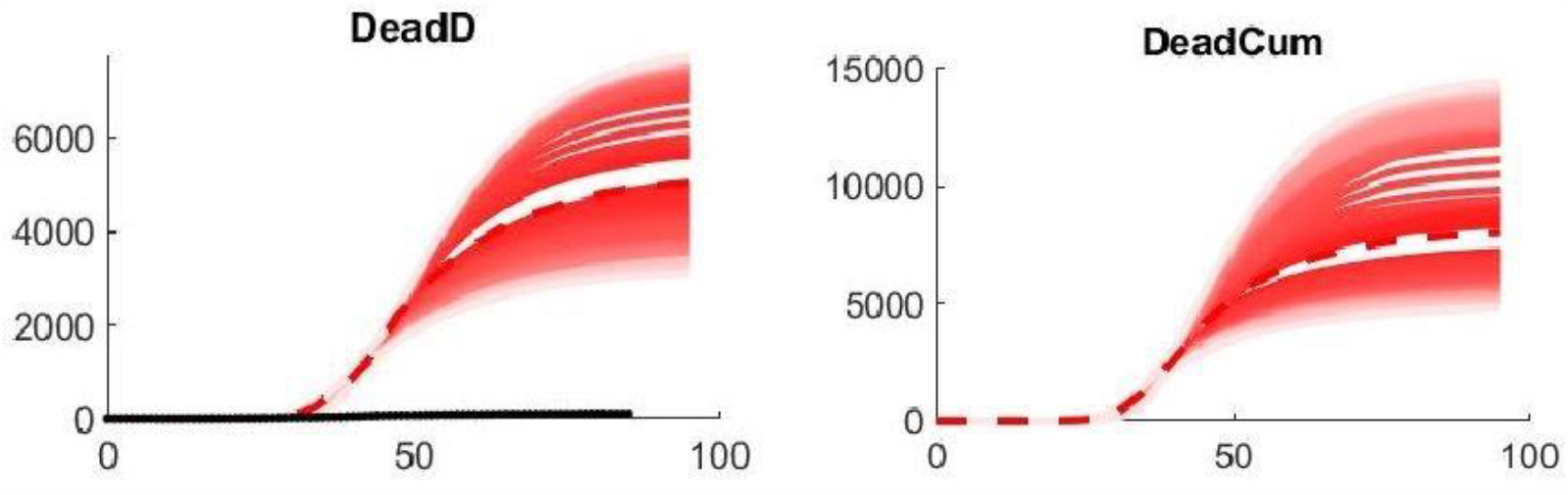
Predicted no lockdown at all scenario. Predicted detected number of deaths (DeadD) and predicted overall deaths (DeadCum) in days if no lockdown or any other measure applied at all. Legend as in Figure 2.

**Figure 4:**
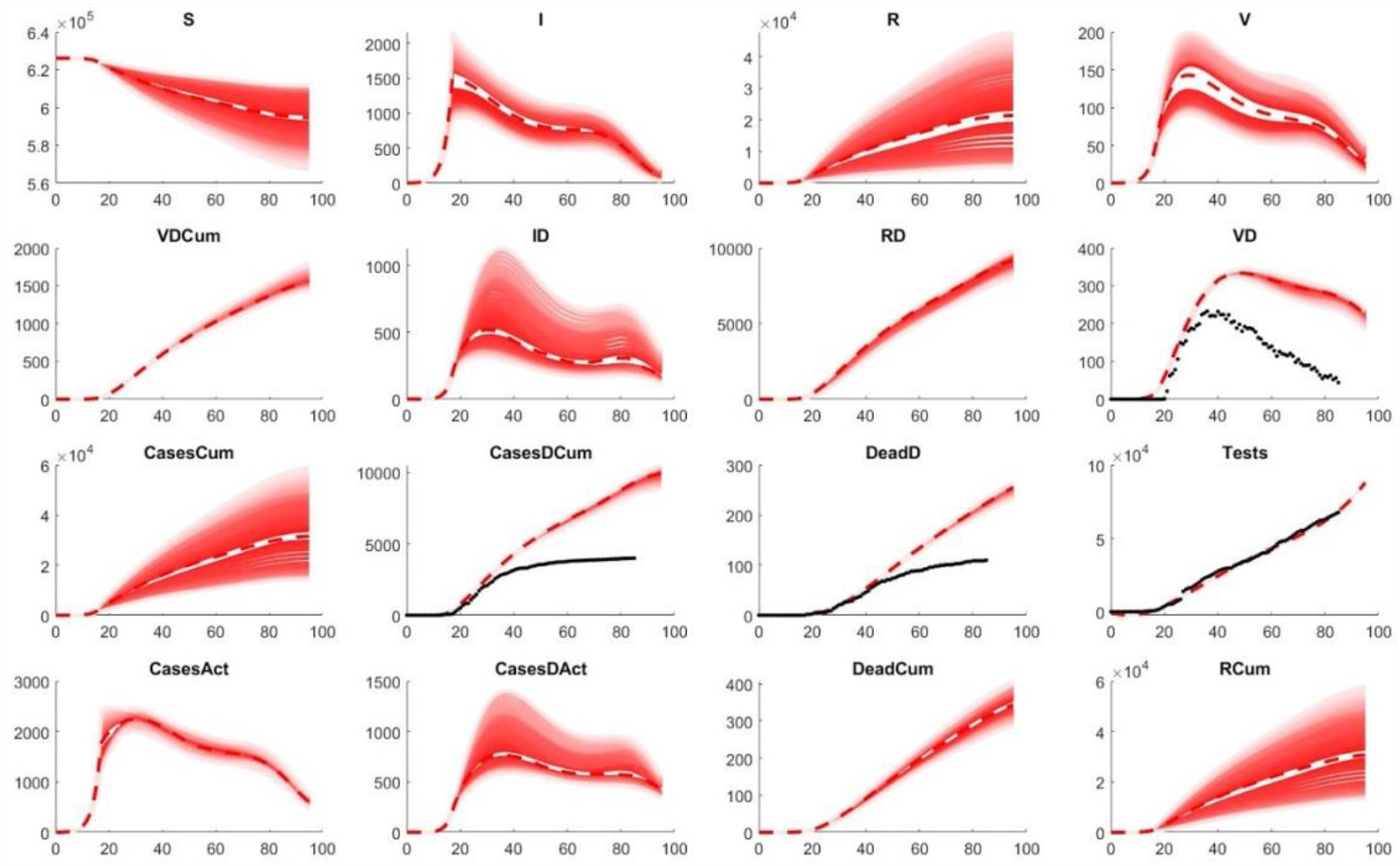
Predicted light lockdown scenario. Example simulation showing a risk of second infection wave and a higher number of cases and deaths, compared to the actual counts (black dots). Legend as in Figure 2.

**Figure 5:**
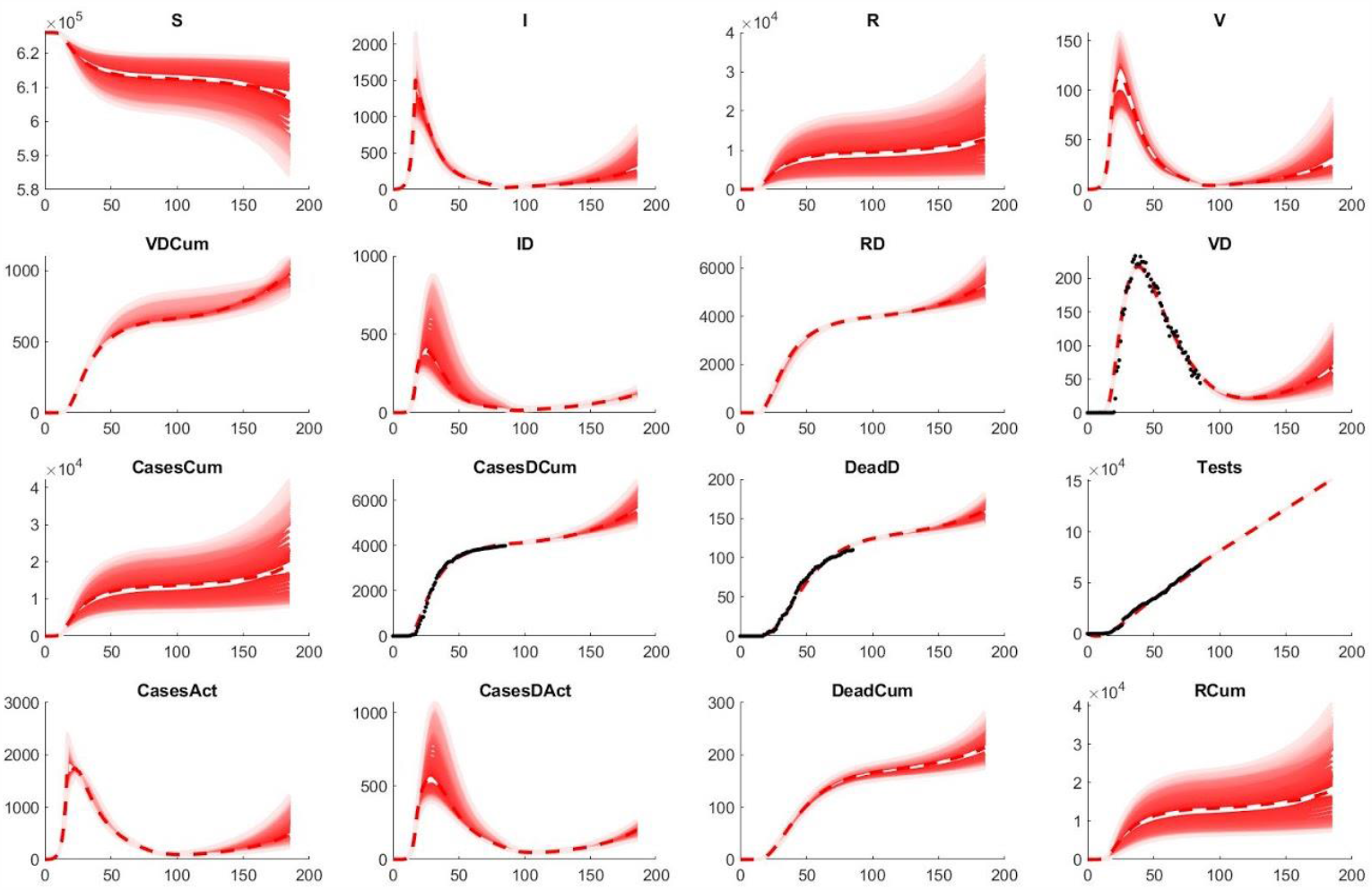
Predicted partial lifting of the lockdown scenario as of end of May (Day 85). Example simulation showing the risk of second infection wave arising around mid of July (Day 135). Legend as in Figure 2.

**Figure 6:**
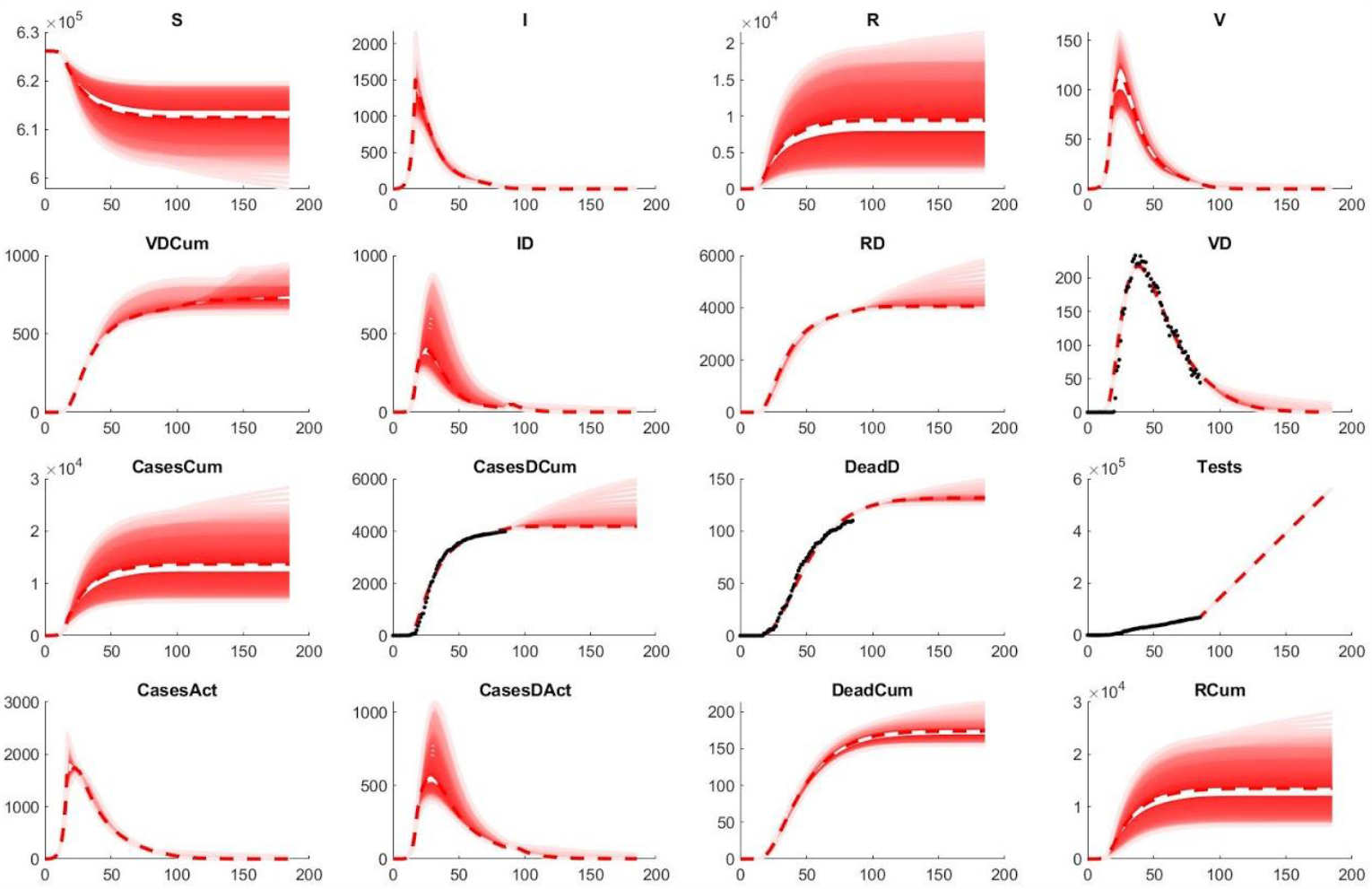
**Predicted lifting of the lockdown scenario as of end of May (Day 85) with increased testing approximately matching the Luxembourgish strategy of testing**. Example simulation showing a reduced risk of second infection wave arising around mid of July (Day 135), compared to lower testing as shown in Figure 5. Legend as in Figure 2.

## RESULTS

As the number of performed tests strongly influences the dynamic analysis of the COVID-19 pandemic in a country or region, we developed a novel *SIR* based epidemiological model (*SIVRT*, Figure 1) which allows the integration of this key information. The model consists of two layers describing the undetected and detected cases whereby the transition between these layers is realized by testing. The model distinguishes severe from non/less symptomatic cases. The probability for severe cases to get tested is assumed to be higher. The model consists of 9 states and has been implemented in the IQM toolbox within Matlab (Methods & Appendix 1). Importantly it allows the fitting to epidemiological data, among others to detected cases, deaths, and currently active cases while staying consistent with the performed tests. The number of tests is impressed on the dynamic model by a polynomial derivative (Methods). Depending on the available data, this allows for a more accurate description of the full (hidden) pandemic dynamics and e.g. allows estimating non-symptomatic cases and the infection fatality rate. The model also allows simulating the effect of partial release of the lockdown (deconfinement) in combination with the planned testing.

In this work, this is exemplified for the development of the first phase of the COVID-19 pandemic in Luxembourg (until end of May 2020). The cumulative numbers of detected cases (*CasesDCum*), the numbers of detected death cases (*DeadD*), as well as the numbers of active hospitalized cases in normal and intense care (as a surrogate for the detected active severe cases, *V*_*D*_) and the cumulative numbers of performed tests (*T*) have been collected for the country in a time-resolved manner (Appendix 2). The model has been fitted to these data by repeated parameter estimation and the successful fits were used for simulation and prediction (Methods). The model nicely fits the observed pandemic in Luxembourg (Figure 2). Furthermore, estimates for the dynamic behavior of the non-observed variables are provided along with confidence intervals derived from the simulations of the successful fits. The rather large confidence intervals for some of the variables indicate that there is not yet enough data to fully identify all parameters of the models. This should be improved in the future by incorporating more data during the course of the ongoing pandemic.

Based on the current modelling, the number of overall cases (*CasesCum*) including detected and non-detected cases as of end of May 2020 is estimated to approximately 13.000 with a large confidence interval (7.000 - 25.000). With an overall population in Luxembourg of 626.108, this gives an estimate of around 2,1% of the population (1,1 - 4%). This estimate matches the result of the CON-VINCE study which indicated a population-wide infection in 1,9% of the Luxembourgish population in antibody-based testing (Snoeck et al., 2020). As mentioned the results of this study and similar studies in other countries can be used to further consolidate the model and improve its accuracy and predictive power.

These model simulations can also be analyzed in terms of the infection fatality rate (IFR, deaths per infection) and the effective reproduction number *Rt_eff*. The model-based estimates of the IFR are around 1,3% with 13.000 estimated cases and 165 estimated overall deaths. This is in agreement with the Case Fatality Rate (CFR, a surrogate of the IFR) in Korea as calculated on March 25, 2020, which had one of the largest numbers of cases and tests performed at the onset of the pandemic (Kim et al., 2020) and more importantly with the estimated IFR after adjusting for delay from confirmation to death obtained on the Diamond Princess Cruise Ship (Russell et al., 2020). The average effective reproduction number *Rt_eff* for Luxembourg can be derived from model parameter kSI. Based on the parameter distribution of the successful model fits, it is estimated to 0.94 (0,70 – 1,17) which is consistent with the average of the respective estimates of the Luxembourgish COVID 19 Research Taskforce (https://researchluxembourg.lu/publications/).

Besides fitting to and analyzing existing data, the model can also be used to predict and analyze alternative scenarios for past and future strategies. For example, when simulating what would have happened in the past until now, if the lockdown would not have taken place at all and the same number of tests would have been performed, a total number of deaths of around 8.000 are predicted to have occurred in Luxembourg with only around 5.000 deaths being detected and assigned to the pandemic (Figure 3). As the same number of tests in this simulation was assumed as performed in reality, a high number of deaths would not have been detected. Whereas, a lighter lockdown could have led already to a second infection wave as shown in an example simulation (Figure 4). Thus obviously also the model supports the huge necessity of the performed lockdown, in comparison with alternative scenarios.

Furthermore, based on the model trained with the data as of end of May, predictions for future scenarios can be performed, especially focusing on the (partial) lifting of the lockdown. A partial lifting of the lockdown can thereby result in a renewal of active cases within around 50 days (Figure 5). This matches the timeframe of the rise of cases which is currently seen in Luxembourg as of mid of July. With increased testing as now performed in Luxembourg, this second wave is predicted to stay small compared to the first wave (Figure 6). Adjustments of the infectability of detected cases might be necessary in future model version to quantitatively describe this second wave.

In summary, the novel testing informed *SIVRT* model structure allows to describe and analyze the COVID-19 pandemic data of Luxembourg in dependency of the number of performed tests. This enables the estimation of the overall and recovered cases, including detected and non-detected cases and thereby the estimation of the infection fatality rate (IFR). It is furthermore possible to perform predictions on past and future scenarios of combinations of lockdown lifting and testing.

## DISCUSSION

Simulations of the novel *SIVRT* model with parameters estimated from the data of the early pandemic in Luxembourg give a full dynamic picture including detected and non-detected cases. In particular, the overall number of cases until end of May and the IFR is estimated at around 13.000, representing 2,1% of the population and 1,3%, respectively. This is in line with 1,9% of volunteers in the CON-VINCE study that had IgG antibodies against SARS-CoV-2 in their plasma and the estimated IFR rate on the Diamond Princess cruise ship 1,3 (95% CI: 0.38–3.6) (Russell et al., 2020). The SIVRT model also allowed predicting the appearance of a second wave in a time frame of 50 days after a partial lifting of the lockdown. This is in concordance with the rise in cases see in Luxembourg as of mid of July.

Although this first case study demonstrates that the novel *SIVRT* model can capture the pandemic dynamics in a country while being informed by the number of performed tests, there are still uncertainties on some of the model states, estimated parameters, and numbers. As next steps, more information should be included to improve the reliability and predictivity of the model. This could include data from earlier and ongoing population-wide screening which gives estimates of overall cases at specific time points. A model-based integration of such information as demonstrated here might especially be useful in countries where population-wide screening is rarely possible or not possible at all, e.g. due to financial constraints. Furthermore, data on recovered and active cases could be incorporated if available in sufficient quality and consistency, as well as information on the sensitivity and specificity of the applied tests. Activity data like travel information could be used to better inform the model parameters describing the different lockdown and deconfinement phases. This would allow to more accurately simulating such scenarios.

Uncertainty in the model could also arise from the data itself. Data on Recovered people is often of poor quality. Obviously undiagnosed cases are not included in the recovered statistics. But, even for detected cases, little attention was given to the number of Recovered. Even, the criteria for recovery vary across countries. After a period of quarantine of 14 days and one or two days without symptoms, patients are assumed to be healed in some countries whereas for others a negative PCR test is required. In Luxembourg, at the onset of the pandemic, the number of recoveries stagnated at 500 individuals over a long period. Only later refinement of the numbers was issued. The number of Recovered impacts directly the number of Susceptible in the model, which is equal to the total population minus all the other states, and hence both states display the largest standard deviation. Another source of uncertainty concerns the testing. In Luxembourg, the number of tests reflects the samples processed and not the individuals tested, and hence negative cases might be overrepresented as individuals might get tested several times.

In follow up work and to assess the general applicability of the model, it should be applied to different countries with different pandemic dynamics and different testing strategies. This would allow deriving country-specific numbers of undetected cases in dependency of the applied testing strategies and assess if key pandemic properties like the IFR or *RT_eff* are comparable between countries. Regularization approaches applied during parameter estimation could thereby help to consolidate significant similarities and differences among countries or regions.

Finally, the model could be extended to include the age structure of the population or the geographic repartition of the cases. However this data is often only available later during the pandemic or for few time points, which limits its usefulness. Furthermore, increasing the number of states does not necessarily improve the prediction power of the model as more data will then be required to parameterize it. In general, simpler models should be preferred over complex ones, as at the onset of the epidemic, when the models are most needed, not enough information of sufficient quality is available and hence unknown parameters have to be obtained by model fitting, which increases the uncertainty of the model (Roda et al., 2020). However, these simpler models should still allow capturing the biological realism of the epidemic, which *SIR* and *SEIR* models often fail to do. The lack of integrated testing information could result in an inconsistency between i.e. the number of Infected described by the model and the respective available data. Hence without a correction for different testing, model predictions would yield very different IFR and *Rt_eff* for each country, resulting in a reduced general applicability of such models. The presented testing informed *SIVRT* model overcomes this issue and shows good predictive potential in this case study for Luxembourg. It thus might is a promising tool for assessing the dynamics of the COVID-19 pandemic by incorporating the number of performed tests.

## Data Availability

For Luxembourg data on people tested positive, death cases, hospitalisation and performed tests were obtained from the website of the Luxembourgish government https://coronavirus.gouvernement.lu/en.html and are summarized in Appendix 2 of this paper.

## APPENDIX Appendix 1

SIVRT model for Luxembourg in IQM toolbox format, resembling the testing data as of May 25, 2020. A derivative of the polynomial fit of the number of tests per day is contained in the *derf* model function. Parameter values are a representative set of a successful model fit.

********** MODEL NAME

SIVRT

********** MODEL NOTES

SIVRT - more detailed and extended for testing

T. Sauter, from 3.4.2020 on, COVID-19 period

********** MODEL STATES

d/dt(S) = -rSI

d/dt(I) = +rSI -rIR -rIV -rIID

d/dt(R) = +rIR +rVR

d/dt(V) = +rIV -rVR -rVD -rVVD

d/dt(Dead) = +rVD

d/dt(ID)= +rIID -rIDVD -rIDR

d/dt(RD) = +rIDR +rVDR

d/dt(VD)= +rVVD +rIDVD -rVDR -rVDD

d/dt(DeadD) = +rVDD

d/dt(CasesCum) = +rSI d/dt(CasesDCum) = +rIID +rVVD

d/dt(VDCum)= +rVVD +rIDVD

d/dt(Tests) = +rVVD +rIID +rSRCumSRCum

S(0) = 626108

I(0) = 1

R(0) = 0

V(0) = 0

Dead(0) = 0

ID(0) = 0

RD(0) = 0

VD(0) = 0

DeadD(0) = 0

CasesCum(0) = 0

CasesDCum(0) = 0

VDCum(0) = 0

Tests(0) = 0

********** MODEL PARAMETERS

kSI = 1.2711

kmSI = 187807

kIR = 0.5249

kIV = 0.02

kVR = 0.0482

kVD = 0.0123

kVVD = 0.1065

kTIvsS = 70.36

kSIld = 0.5260

ld = 0

ldday = 17

********** MODEL VARIABLES

kIDR = kIR

kIDVD = kIV kVDR = kVR kVDD = kVD

CasesAct = I + ID + V + VD

CasesDAct = ID + VD

RCum = R + RD

DeadCum = DeadD + Dead

********** MODEL REACTIONS

derTests = derf(time)

rSI = kSI*(1+ld*(kSIld-1))*(S/(kmSI+S))*(CasesAct-ID-VD)

rIR = kIR*I

rIV = kIV*I

rVR = kVR*V

rVD = kVD*V

rVVD = kVVD*V

rIID = (derTests-rVVD)*kTIvsS*I/(S+kTIvsS*I+RCum)

rIDVD = kIDVD*ID

rIDR = kIDR*ID rVDR = kVDR*VD

rVDD = kVDD*VD

rSRCumSRCum = (derTests-rVVD)*(S+RCum)/(S+kTIvsS*I+RCum)

********** MODEL FUNCTIONS

derf(a) =0.020527*a^3-2.9703*a^2+130.7401*a-747.5901

********** MODEL EVENTS event = gt(time,ldday),ld,1

********** MODEL MATLAB FUNCTIONS

## Appendix 2

Development of the COVID pandemic in Luxembourg indicating the cumulative numbers of detected cases (*CasesDCum*), the numbers of detected death cases (*DeadD*), as well as the numbers of active hospitalized cases in normal and intense care (as a surrogate for the detected active severe cases, *V*_*D*_) and the cumulative numbers of performed tests (*T*).

| Date | *time* | *CasesDCum* | *DeadD* | *VD* | *Tests* |
| --- | --- | --- | --- | --- | --- |
| 28/02/2020 | 0 | 1 | 0 | 0 | 19 |
| 29/02/2020 | 1 | 1 | 0 | 0 | 20 |
| 01/03/2020 | 2 | 1 | 0 | 0 | 20 |
| 02/03/2020 | 3 | 1 | 0 | 0 | 27 |
| 03/03/2020 | 4 | 1 | 0 | 0 | 31 |
| 04/03/2020 | 5 | 1 | 0 | 0 | 31 |
| 05/03/2020 | 6 | 2 | 0 | 0 | 37 |
| 06/03/2020 | 7 | 3 | 0 | 0 | 46 |
| 07/03/2020 | 8 | 4 | 0 | 0 | 52 |
| 08/03/2020 | 9 | 4 | 0 | 0 | 52 |
| 09/03/2020 | 10 | 4 | 0 | 0 | 68 |
| 10/03/2020 | 11 | 4 | 0 | 0 | 93 |
| 11/03/2020 | 12 | 6 | 0 | 0 | 144 |
| 12/03/2020 | 13 | 21 | 0 | 0 | 344 |
| 13/03/2020 | 14 | 45 | 0 | 0 | 547 |
| 14/03/2020 | 15 | 66 | 0 | 0 | 654 |
| 15/03/2020 | 16 | 66 | 0 | 0 | 678 |
| 16/03/2020 | 17 | 99 | 0 | 0 | 1098 |
| 17/03/2020 | 18 | 221 | 2 | 0 | 1748 |
| 18/03/2020 | 19 | 379 | 2 | 0 | 2698 |
| 19/03/2020 | 20 | 522 | 3 | 1 | 3669 |
| 20/03/2020 | 21 | 697 | 5 | 21 | 4777 |
| 21/03/2020 | 22 | 824 | 6 | 62 | 5259 |
| 22/03/2020 | 23 | 845 | 6 | 68 | 5280 |
| 23/03/2020 | 24 | 1087 | 6 | 78 | 6477 |
| 24/03/2020 | 25 | 1281 | 6 | 106 | 7454 |
| 25/03/2020 | 26 | 1451 | 8 | 149 | 8441 |
| 26/03/2020 | 27 | 1714 | 9 | 146 | 13926 |
| 27/03/2020 | 28 | 1910 | 14 | 154 | 15164 |
| 28/03/2020 | 29 | 2037 | 17 | 181 | 16136 |
| 29/03/2020 | 30 | 2081 | 21 | 187 | 16457 |
| 30/03/2020 | 31 | 2252 | 22 | 186 | 17670 |
| 31/03/2020 | 32 | 2364 | 26 | 195 | 18784 |
| 01/04/2020 | 33 | 2555 | 27 | 199 | 20194 |
| 02/04/2020 | 34 | 2684 | 28 | 224 | 21618 |
| 03/04/2020 | 35 | 2801 | 29 | 228 | 22961 |
| 04/04/2020 | 36 | 2872 | 34 | 233 | 23835 |
| 05/04/2020 | 37 | 2911 | 36 | 220 | 24455 |
| 06/04/2020 | 38 | 2988 | 38 | 221 | 25483 |
| 07/04/2020 | 39 | 3075 | 43 | 220 | 26613 |
| 08/04/2020 | 40 | 3155 | 46 | 232 | 27624 |
| 09/04/2020 | 41 | 3226 | 49 | 224 | 28498 |
| 10/04/2020 | 42 | 3274 | 55 | 224 | 29186 |
| 11/04/2020 | 43 | 3286 | 58 | 212 | 29521 |
| 12/04/2020 | 44 | 3305 | 62 | 223 | 29737 |
| 13/04/2020 | 45 | 3308 | 65 | 203 | 29795 |
| 14/04/2020 | 46 | 3383 | 68 | 206 | 31237 |
| 15/04/2020 | 47 | 3433 | 68 | 188 | 32080 |
| 16/04/2020 | 48 | 3477 | 71 | 198 | 32967 |
| 17/04/2020 | 49 | 3532 | 71 | 180 | 33923 |
| 18/04/2020 | 50 | 3551 | 74 | 192 | 34438 |
| 19/04/2020 | 51 | 3559 | 75 | 187 | 34574 |
| 20/04/2020 | 52 | 3590 | 78 | 188 | 35541 |
| 21/04/2020 | 53 | 3638 | 80 | 185 | 36399 |
| 22/04/2020 | 54 | 3656 | 81 | 178 | 37530 |
| 23/04/2020 | 55 | 3676 | 84 | 163 | 38246 |
| 24/04/2020 | 56 | 3708 | 85 | 159 | 39471 |
| 25/04/2020 | 57 | 3723 | 88 | 147 | 40069 |
| 26/04/2020 | 58 | 3729 | 88 | 146 | 40170 |
| 27/04/2020 | 59 | 3741 | 89 | 137 | 40867 |
| 28/04/2020 | 60 | 3774 | 89 | 140 | 42489 |
| 29/04/2020 | 61 | 3790 | 90 | 128 | 44197 |
| 30/04/2020 | 62 | 3804 | 92 | 114 | 46047 |
| 01/05/2020 | 63 | 3811 | 93 | 126 | 47482 |
| 02/05/2020 | 64 | 3823 | 96 | 120 | 48118 |
| 03/05/2020 | 65 | 3827 | 96 | 118 | 48339 |
| 04/05/2020 | 66 | 3839 | 98 | 115 | 49307 |
| 05/05/2020 | 67 | 3850 | 98 | 121 | 50543 |
| 06/05/2020 | 68 | 3859 | 100 | 114 | 51883 |
| 07/05/2020 | 69 | 3871 | 100 | 98 | 53315 |
| 08/05/2020 | 70 | 3877 | 101 | 105 | 54553 |
| 09/05/2020 | 71 | 3886 | 101 | 95 | 55246 |
| 10/05/2020 | 72 | 3888 | 101 | 89 | 55411 |
| 11/05/2020 | 73 | 3894 | 102 | 89 | 56390 |
| 12/05/2020 | 74 | 3904 | 103 | 90 | 57687 |
| 13/05/2020 | 75 | 3916 | 103 | 79 | 58835 |
| 14/05/2020 | 76 | 3923 | 104 | 83 | 59988 |
| 15/05/2020 | 77 | 3932 | 106 | 74 | 61207 |
| 16/05/2020 | 78 | 3945 | 107 | 64 | 62154 |
| 17/05/2020 | 79 | 3947 | 107 | 55 | 62260 |
| 18/05/2020 | 80 | 3959 | 109 | 61 | 63084 |
| 19/05/2020 | 81 | 3971 | 109 | 58 | 64019 |
| 20/05/2020 | 82 | 3980 | 109 | 63 | 65078 |
| 21/05/2020 | 83 | 3981 | 109 | 52 | 65830 |
| 22/05/2020 | 84 | 3990 | 109 | 56 | 66805 |
| 23/05/2020 | 85 | 3992 | 110 | 44 | 67729 |

